# From survey to support: Exploring hospitalized patients’ mental well-being and the opportunities for intelligent assistive devices

**DOI:** 10.1101/2025.08.01.25332768

**Authors:** Annalena Meyer, Nils Rathmann, Marcus Vetter

## Abstract

**Background:** Intelligent assistive technologies, including chatbots and wearable biosensors, hold considerable promise for transforming mental well-being support in healthcare settings. Hospital stays are often associated with emotional strain, influenced by environmental, procedural, and communication factors [1, 2]. Intelligent assistive technologies have the potential to provide real-time, personalized interventions. However, for a successful translation of these technologies into clinical practice, patient insights are imperative. [3–5]

**Methods:** A 2023 cross-sectional survey at the Department of Radiology and Nuclear Medicine at the University Hospital Mannheim aimed to inform the development of intelligent assistive technologies for supporting the patients’ emotional well-being. The survey, which was designed to assess emotional states, psychological stressors, and care experiences, employed a mixed-methods approach, incorporating 39 questions. The data was analyzed using descriptive and inferential statistics, with qualitative responses coded for thematic patterns.

**Results:** The survey revealed that prolonged waiting times (*>* 1 hour, 74% of participants) and communication problems significantly reduced mental well-being (mean score dropped from 4.45 to 3.57 out of 5, *p* = 0.027). Language barriers and organizational inefficiencies exacerbated distress, while staff empathy and social visits improved well-being. Prior to procedures, the most common stated emotions by participants were apprehension (68% therapeutic procedure and 58% diagnostic procedure) and anxiety (47% therapeutic, 21% diagnostic), with these emotions shifting to confidence (78% therapeutic, 57% diagnostic) and hope (62% therapeutic, 49% diagnostic) post-procedure.

**Conclusion:** These findings highlight communication delays and procedural anxiety as key emotional stressors. The strategic implementation of intelligent assistive technologies, such as multilingual AI chatbots capable of providing real-time updates and biosensors monitoring physiological stress, has the potential to mitigate distress and enhance patient experiences, especially in situations with heightened emotional states (eg. pre-procedural phases, extended waiting periods, or limited information flow). Future development should prioritize co-design with patients and clinicians, with effectiveness validated through multi-center clinical trials.

## Introduction

An intelligent assistive system designed to support the emotional well-being of hospitalized patients is being developed. To inform the design of the system, a patient-centered survey was conducted to identify key psychological stressors and perceptions of care within a hospital environment.

When situated within the broader context, hospitalization is a critical period often associated with heightened psychological vulnerability, particularly among pediatric patients and their families. Existing research demonstrates that hospital stays frequently induce stress, anxiety, and psychological distress due to unfamiliar environments, uncertainty regarding outcomes, and disruptions in daily routines [1, 2, 6]. Addressing these challenges has become a growing priority in healthcare, with increasing emphasis on interdisciplinary approaches that combine clinical insight with technological innovation to improve patient experiences and outcomes [7, 8].

Digital assistive technologies are emerging as valuable tools in this context, offering scalable and cost-effective solutions to support patients’ mental health during hospitalization. Mobile health applications and chatbot systems provide psychoeducational support, self-management tools, and therapeutic engagement, fostering emotional resilience and patient autonomy [3, 9]. Coinciding, healthcare providers benefit from decision support tools and diagnostic aids that enhance clinical workflow, accuracy, and ultimately, patient outcomes [10–12].

The application of assistive technologies extends beyond mental health to address a broad spectrum of medical conditions. Cognitive training platforms and wearable devices, for instance, have demonstrated potential in managing attention deficit hyperactivity disorder (ADHD) by improving focus and reducing impulsivity [13]. In the context of chronic disease management, digital behavioral interventions have demonstrated efficacy in alleviating depressive symptoms among patients with Type 1 and Type 2 diabetes mellitus [14]. Furthermore, advancements in computer vision support diagnostic imaging, patient safety through fall detection, and respiratory surveillance through non-invasive monitoring [15]. Despite the optimism these advancements may inspire, many authors underscore the necessity for additional empirical validation to ensure reliability, efficacy, and ethical integration into clinical practice. [14, 16]

Recent advancements in digital health technologies provide promising solutions to mitigate these challenges. Intelligent assistive devices, mobile applications, and AI-driven chatbots have demonstrated potential in delivering real-time updates, personalized coping strategies, and automated counseling support [5, 17–19]. Notably, research suggests that digital platforms can enhance communication between patients and healthcare providers, reducing uncertainty and stress [4, 14]. Implementing such tools in clinical settings could foster a more patient-centered approach, addressing both logistical inefficiencies and psychological burdens associated with hospitalization. However, their effectiveness remains an area for further investigation, particularly regarding their adaptability across diverse patient populations and hospital infrastructures.

Despite growing interest and innovation in digital health, gaps remain in understanding the specific factors that influence emotional well-being during hospitalization, particularly from the patient’s perspective. Extended wait times, limited communication, and unfamiliar settings continue to significantly impair mental well-being. These challenges underscore the necessity of intelligent assistive systems tailored to address the psychological needs of patients in real-time hospital environments.

The objective of the survey was to set the groundwork for an intelligent assistant intended to support patients’ emotional well-being in a hospital setting. To accomplish this objective, the factors that impact mental well-being during hospitalization were examined. Participants were questioned about their personal treatment experiences, including their perceptions of organizational structures, interactions with hospital staff, and their emotional states. The survey was conducted to confirm and quantify the listed factors for the specific hospital. These results are used as a basis for designing and developing intelligent assistive devices to support the mental well-being of hospitalized patients.

## Materials and methods

The survey was designed as a cross-sectional, observational study. The data collection was carried out between July 3, 2023, and August 18, 2023. The interviews for the survey were performed at the University Hospital Mannheim’s Department of Radiology and Nuclear Medicine.

### Sample and procedure

Utilizing a mixed methods design, the survey gathered data through in-person interviews conducted with a commitment to participant anonymity. Patient recruitment took place in the waiting area of the radiology department, employing a convenience sampling approach. The survey’s inclusion criteria required hospitalization and receiving a radiological procedure. Minors were excluded. The sample comprises 51 interviews with hospitalized patients.

The research project received approval from the Ethics Committee II of the University of Heidelberg (approval number 2023-577). The committee waived the requirement for informed consent. Participants did not receive financial or other incentives.

### Measures

In order to facilitate participation by all patients, irrespective of their physical conditions, the survey was conducted in person. Conversations were deliberately kept short and focused to optimize completion rates. The interviews were only carried out during waiting periods to avoid interference with the required care and treatment. The patients in the waiting area were situated in beds or wheelchairs. To maintain a high completion rate, the survey was designed with a minimal number of questions in simple language. In addition to closed-ended questions with predetermined answer options, open-ended questions were incorporated. These open-ended queries aimed to capture nuanced details and factors that might not have been covered by the closed-ended questions, providing a comprehensive representation of the participants’ experiences and insights. [20]

### Data analysis

The analytical phase utilized both descriptive and inferential statistical methods, coupled with an intuitive coding strategy customized for open-ended questions. Descriptive statistics were used to provide an overview of the acquired data, including measures of central tendency and variability. Meanwhile, inferential statistical tests were leveraged to draw broader insights from the dataset. The CROSS method [21] was applied for reporting. Demographic questions were strategically included in the survey to discern potential disparities in mental well-being among various participant groups during their hospital stay. Wilcoxon signed-rank analysis was used to examine potential variations. Correlation analysis was conducted to identify complex patterns, trends, and relationships between various observations within the dataset. Sensitivity analyses, including nonparametric group comparisons, were conducted to assess the robustness of the findings. In adherence to statistical conventions, a significance level (*α*) of 0.05 was set for all tests. Cases with missing data were excluded pairwise from each statistical test. Table 1 shows an overview of the collected data.

**Table 1.** Comprehensive Survey Data on Hospitalized Patients’ Mental Well-Being.

| Characteristic (RR %) | Category | n (%) |
| --- | --- | --- |
| **Demographics** |  |  |
| Gender (100.0%) | Male | 42 (82.4%) |
|  | Female | 9 (17.6%) |
| Age Group (98.0%) | 18-34 | 4 (8.0%) |
|  | 35-44 | 3 (6.0%) |
|  | 45-54 | 4 (8.0%) |
|  | 55-64 | 10 (20.0%) |
|  | 65-74 | 17 (34.0%) |
|  | 75-84 | 9 (18.0%) |
|  | 85+ | 3 (6.0%) |
| **Hospital Stay** |  |  |
| Planned Procedure (96.1%) | Diagnostic | 30 (61.2%) |
|  | Therapeutic | 19 (38.8%) |
| Intervention Conducted (90.2%) | Yes | 18 (39.1%) |
|  | No | 28 (60.9%) |
| First Procedure (98.0%) | Yes | 16 (32.0%) |
|  | No, procedure 2 or 3 | 19 (38.0%) |
|  | No, procedure 4+ | 15 (30.0%) |
| Hospital Ward (92.2%) | surgery | 13 (27.7%) |
|  | urology | 13 (27.7%) |
|  | oncology | 4 (8.5%) |
|  | nephrology | 3 (6.4%) |
|  | neurology | 3 (6.4%) |
|  | Other wards | 11 (23.4%) |
| First Hospital Stay (98.0%) | Yes | 10 (20.0%) |
|  | No | 40 (80.0%) |
| Previous Stay at UMM (74.5%) | Yes | 31 (81.6%) |
|  | No | 7 (18.4%) |
| Planned Stay Duration (82.4%) | <2 days | 15 (35.7%) |
|  | 3-5 days | 8 (19.1%) |
|  | 5-7 days | 12 (28.6%) |
|  | 8-10 days | 2 (4.8%) |
|  | >10 days | 5 (11.9%) |
| Stay Postponed (82.4%) | Yes | 13 (31.0%) |
|  | No | 29 (69.0%) |
| **Social Factors** |  |  |
| Regular Visitors (96.1%) | Family | 43 (87.8%) |
|  | Friends | 24 (50.0%) |
|  | No | 6 (12.2%) |
| Regular Contact (98.0%) | Family | 50 (100.0%) |
|  | Friends | 42 (84.0%) |
|  | No | 0 (0.0%) |
| Feeling isolated (88.2%) | Yes | 4 (8.9%) |
|  | No | 41 (91.1%) |
| Communication satisfaction (94.1%) |  | 4.54 ± 0.82 |
| **Waiting Times** |  |  |
| Waiting Times (92.2%) | <30 min | 8 (17.0%) |
|  | 30 min - 1h | 5 (10.6%) |
|  | 1-2h | 14 (29.8%) |
|  | >2h | 20 (42.6%) |
| Perceived Wait (90.2%) | Short | 4 (8.7%) |
|  | Neutral | 18 (39.1%) |
|  | Long | 24 (52.2%) |
| Waiting Activities (88.2%) | Lying in bed | 7 (15.6%) |
|  | Phone | 6 (13.3%) |
|  | Other | 32 (71.1%) |
| Waiting time Impact on<br>mental well-being (88.2%) | Yes | 16 (35.6%) |
|  | No | 29 (64.4%) |
| **Emotional States** |  |  |
| Pain level (86.3%) |  | 1.88 ± 1.77 |
| Emotions Before (88.2%) | anxious | 14 (31.1%) |
|  | apprehension | 28 (62.22%) |
|  | confused | 1 (1.0%) |
|  | happy | 1 (1.0%) |
|  | stressed | 6 (13.3%) |
|  | confident | 20 (44.4%) |
|  | hopeful | 17 (37.8%) |
|  | unsure | 2 (1.9%) |
|  | neutral | 11 (24.4%) |
|  | other | 4 (8.9%) |
| Emotions After (31.4%) | anxious | 1 (3.2%) |
|  | apprehension | 2 (6.5%) |
|  | confused | 0 (0.0%) |
|  | happy | 6 (37.5%) |
|  | stressed | 0 (0.0%) |
|  | confident | 11 (68.8%) |
|  | hopeful | 7 (43.75%) |
|  | unsure | 1 (6.3%) |
|  | neutral | 1 (6.3%) |
|  | other | 2 (12.5%) |

The analytical processes were executed using Python 3.11 using pandas, scipy, statsmodels and miceforest libraries.

## Results

### Sample description

The interviews were conducted with hospitalized patients between July and August 2023. Of the sample 18% were female (cf Tab 1). The majority of participants (80%) had experienced prior hospitalizations. Of all participants, 61% had been admitted to the University Hospital Mannheim before. A significant proportion of participants (61%) underwent diagnostic imaging, such as CT scans, and the remaining 39% of patients underwent minimally invasive procedures (e.g., angiographic procedures, CT-guided needle interventions).

### Mental well-being

Mean self-reported mental well-being, measured on a 5-point scale, decreased from 4.45 (*σ*=0.91) prior to hospitalization to 3.57 (*σ*=1.10) during hospitalization, highlighting the emotional toll of inpatient care, as shown in Fig 1. Women stated higher scores for well-being at home and lower scores as in-patients (at home 5 and *σ* = .00, hospitalized 3 and *σ* = 0.96; p = .2). However, no statement about the significance can be made because the number of participating women (N = 8) is too small. The predominant response among the participants who reported changes in their well-being was “not being at home” (16 out of 40). However, a small number of participants reported experiencing improved well-being after leaving their domestic setting. These exceptional cases were associated with factors such as feelings of loneliness in the domestic environment, the recent bereavement of a family member, or the receipt of assistance and a renewed perspective during a period of hospitalization.

**Fig 1.**
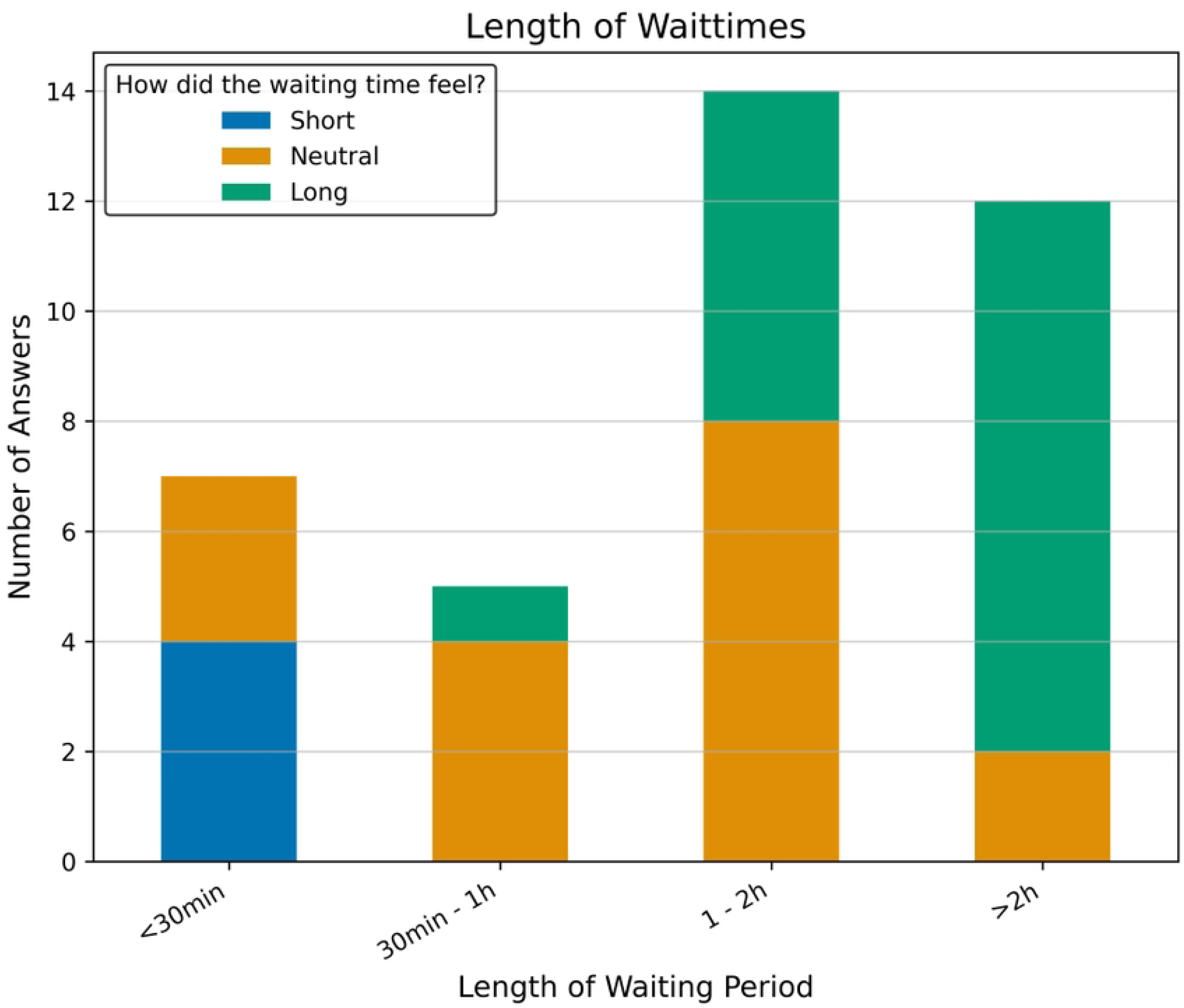
Mental well-being scores before and during hospitalization. The box plots show a decrease in well-being scores during hospitalization. The shaded boxes represent the interquartile range, whiskers indicate data spread, outliers are shown as open circles, and the mean is marked by an orange dot.

A moderate inverse correlation was observed between the planned length of hospitalization and reported well-being scores (*ρ* = –0.41, p = 0.027), indicating that longer expected stays may negatively impact patients’ mental states. Furthermore, the ward of admission was identified as a significant factor influencing well-being (p = 0.034).

Participants were queried regarding the duration of their waiting periods during their hospital stay, with the question presenting predefined time intervals as response options. The majority of participants reported extended waiting times exceeding one hour (74%). In addition to the length of the waiting period, participants were asked to describe how they perceived the waiting period. The presented answer options were “short”, “neutral”, and “long”. The perceived waiting times are strongly correlated with the actual durations (*ρ* = .744, p *<* .001, see Fig 2), underscoring patients’ sensitivity to procedural delays. Among the participants who had waited less than 30 minutes (17%), the majority perceived the waiting time as “short” (8%). The most common coping activities included sleeping or dozing (41%) and using the smartphone (25%). A significant proportion of the participants, 35.6%, reported that prolonged waiting periods had a negative effect on their mental well-being.

**Fig 2.**
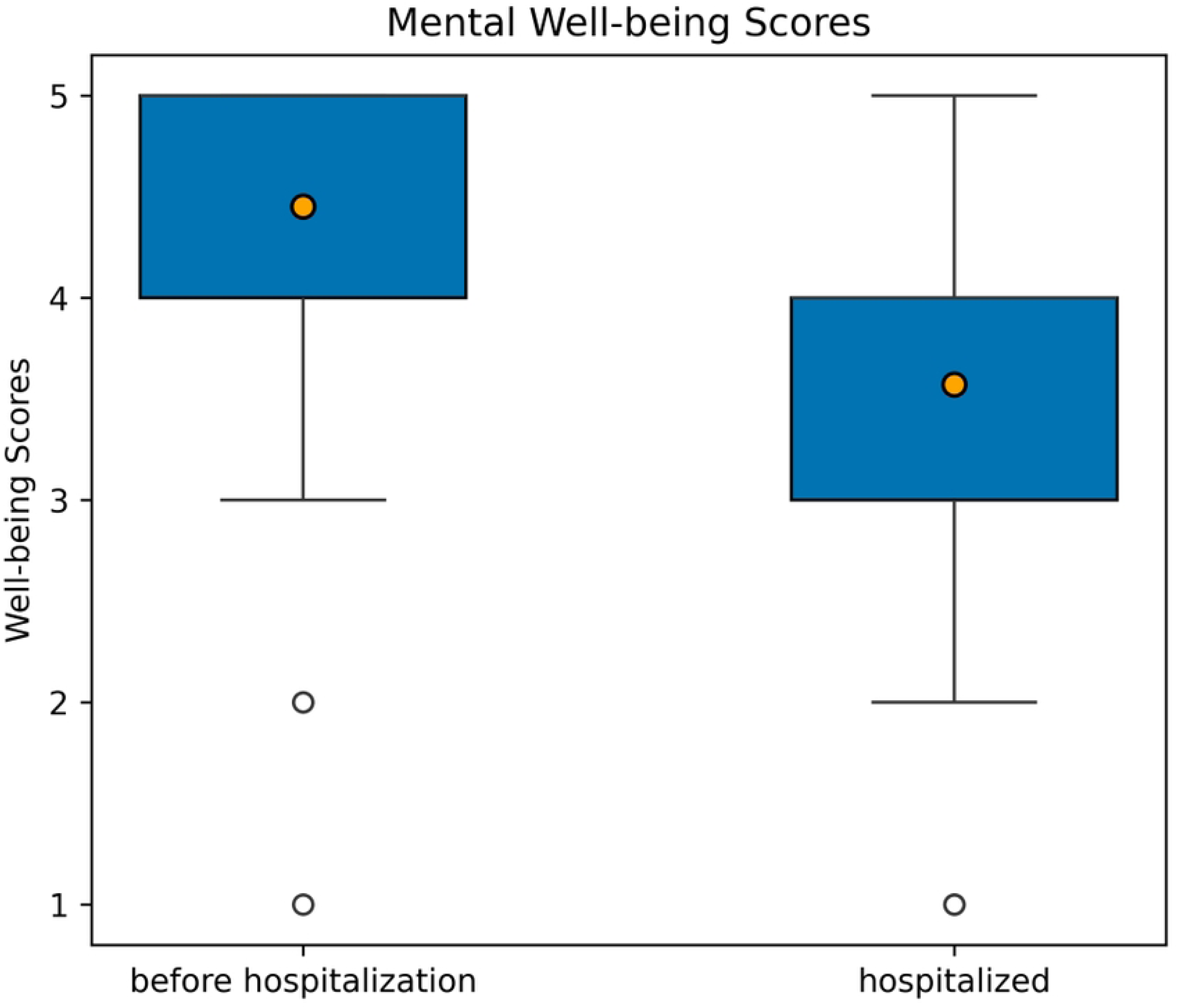
Perceived waiting time in relation to waiting periods. The x-axis represents different waiting time categories: less than 30 minutes, 30 minutes to 1 hour, 1 to 2 hours, and more than 2 hours. The y-axis shows the number of responses. The stacked bars indicate how the waiting time was perceived, categorized as short (blue), neutral (orange), or long (green).

The analysis revealed that the overall satisfaction of the participants with the communication from healthcare professionals was high with a mean of 4.54 (*σ* = .82). No significant gender differences were observed regarding communication (men = 4.5, women = 4.55; p = .71). However, no correlation to the mental well-being was found. The subsequent qualitative responses indicated the presence of several critical stressors that were not measured by quantitative means. It is noteworthy that the most prevalent communication breakdowns transpired during protracted waiting periods, frequently attributable to staff members’ inability to furnish prompt or comprehensive procedural information. These experiences have shown to contribute to emotional distress and feelings of uncertainty. This suggests that infrequent communication was more distressing than the content of conversations itself. Moreover, participants with limited German language proficiency indicated challenges in interacting with clinical staff, particularly due to the lack of consistent English-language support. Multiple participants observed that communication was fragmented across departments, thus indicating systemic organizational deficiencies. These findings underscore the emotional and structural challenges posed by inadequate information flow and language barriers in care settings. They also highlight the potential value of multilingual, AI-driven communication tools in addressing information flow discrepancies, enhancing emotional reassurance, and supporting patient-centered care throughout the hospitalization process.

Further questions were asked about the emotions participants experienced before and after procedures. As shown in Fig. 3, the question provided several different emotions as response options and the opportunity to provide additional information. The emotional responses varied among participants, but some general trends emerged.

**Fig 3.**
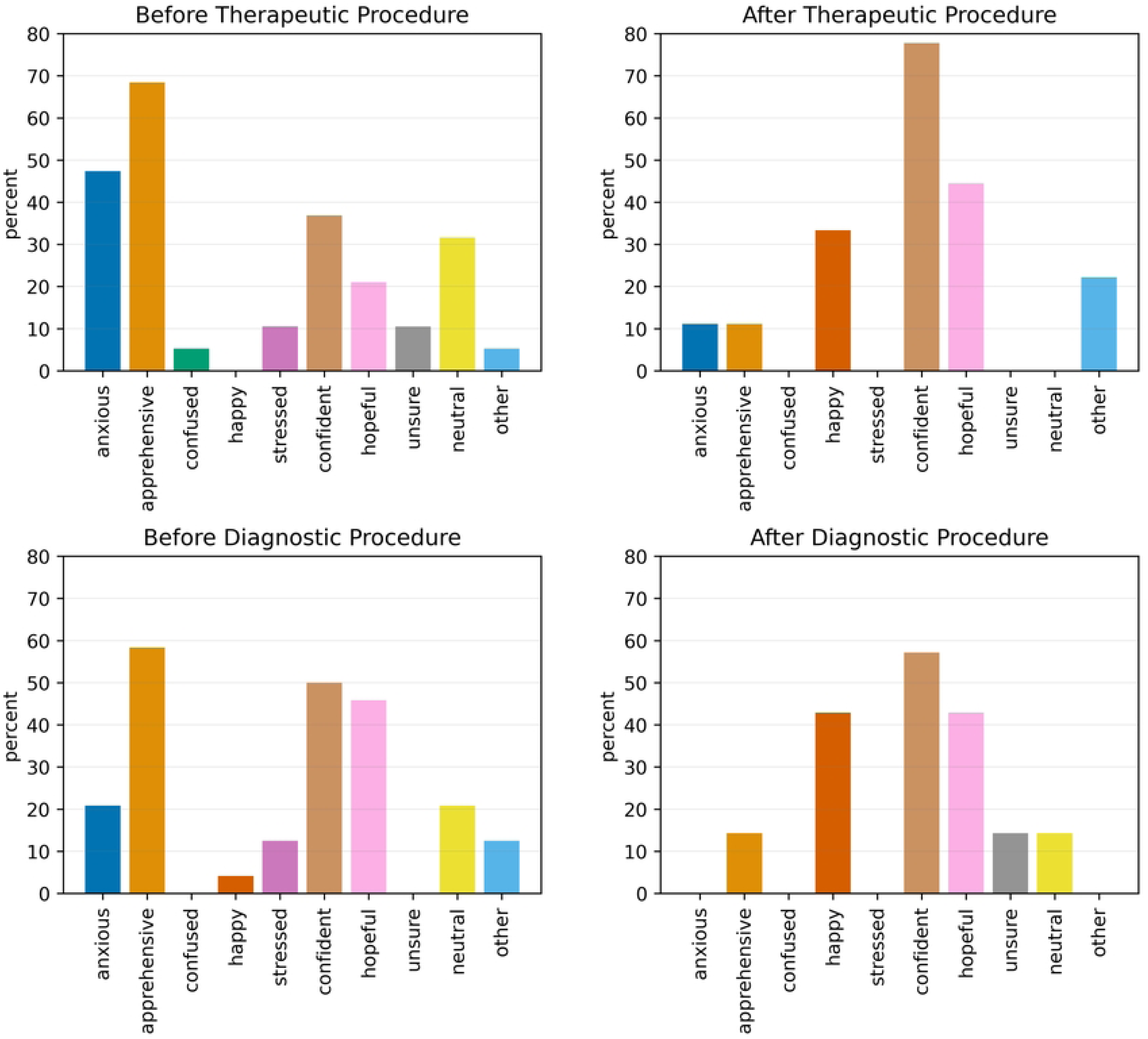
Distribution of emotional states before and after medical procedures. The top row presents data for therapeutic procedures, with the left panel showing the distribution of reported emotions before the procedure and the right panel showing the distribution after the procedure. The bottom row displays data for diagnostic procedures, with the left panel representing emotions before the procedure and the right panel representing emotions after the procedure. The x-axis categorizes different emotional states, while the y-axis indicates the percentage of individuals reporting each emotion.

Before undergoing therapeutic procedures, the most common emotions reported by participants were apprehension (68%), anxiety (47%), and neutrality (32%), indicating a considerable level of concern. However, a subset of participants also expressed feelings of confidence (37%) and hope (21%). Post-procedure, a notable shift in emotional responses was observed. Confidence (78%), hope (44%), and happiness (33%) became the predominant emotions, with apprehension and anxiety markedly decreasing to 11% each. A comparable trend was noted in the survey preceding diagnostic procedures.

Apprehension (58%), confidence (50%), and hope (46%) were the most frequently reported emotions, with anxiety noted by 21% of participants. Subsequent to these diagnostic procedures, emotional responses mirrored those following therapeutic interventions, with confidence (57%), happiness (43%), and hope (43%) being the most prevalent. A striking observation was the complete absence of stress reports after diagnostic procedures, contrasting with the persistence of stress in therapeutic contexts, where it decreased but remained in 11% of cases.

Participants encountered difficulties in identifying specific factors that positively influenced their mental well-being. Some participants offered generalized responses, such as “all okay” (N = 9). Upon the identified specific factors, social and interpersonal influences were most frequently cited, such as “kindness and professionalism of the staff”(N = 16) and “being visited” (N = 8). Conversely, organizational factors, such as “long waiting times” (N = 7) and communication issues (N = 4), were frequently mentioned as negative influences on mental well-being. These findings underscore the importance of developing assistive technologies that not only respond to emotional fluctuations but also address contextual stressors (e.g., waiting times, communication breakdowns, and type of procedure), thereby enhancing the patient experience and promoting overall mental well-being.

A comparative analysis of emotional states between procedure groups was conducted, revealing both similarities and distinctions (see Fig. 3). Before the procedure, a state of apprehension was prevalent, affecting 68% of patients undergoing therapeutic procedures and 58% of those undergoing diagnostic procedures. Anxiety was greater among therapeutic patients at 47% compared to 21% in the diagnostic group. The differences suggest that procedural factors influence patients’ mental well-being. After procedures, both groups shifted to positive emotions, with confidence reported by 78% of therapeutic patients and 57% of diagnostic patients, and hope expressed by 44% and 43% respectively, suggesting comparable relief. Stress persisted in 11% of therapeutic patients post-procedure but was absent in the diagnostic group, suggesting a more persistent mental burden for therapeutic patients. The distinctions show an opportunity for intelligent assistive technologies that are developed to address the needs of patients in these contexts.

Most interviewees reported little to no pain, with an average score of 1.88 (*σ* = 1.77) out of 10 during the survey. A significant association between pain intensity and the presence of anxiety was observed (*ρ* = 0.34, p = .021; U = 132.5, p = .023), suggesting that physical discomfort may contribute to emotional distress.

## Discussion

This survey emphasizes the emotional toll of hospitalization on patients undergoing radiological procedures and underscores the potential of intelligent assistive technologies to support patient-centered care. This investigation offers a distinctive perspective on patient mental well-being, augmenting prior research on patient satisfaction and mental well-being in clinical settings [2, 22–24]. Our survey supports existing research by confirming a decline in mental well-being during hospitalization, often attributed to the unfamiliar environment. With 74% of patients reporting prolonged waiting times and 65% identifying organizational challenges, these factors emerged as significant contributors to psychological distress. These findings are consistent with the consensus that mental well-being is a fundamental aspect of comprehensive patient care [23, 24].

A notable finding is that 73% of the participants reported experiencing emotional fluctuations throughout their hospital stay, transitioning from anticipatory anxiety before procedures to relief post-intervention. This pattern aligns with previous findings on psychological responses to clinical interventions [22, 25, 26], and underscore the clinical relevance of addressing distress, especially in a pre-procedural phase. Intelligent assistive technologies offer scalable solutions to provide adaptable patient support. For instance, AI-powered conversational agents may deliver real-time emotional support and information, helping patients manage mentally distressing moments such as waiting periods or unfamiliar clinical settings [27]. Recent advances, such as the AIME framework from Google Research [19], illustrate how multi-modal systems that integrate speech, visuals, and situational context can interpret patient needs and deliver multilingual, emotionally responsive communication. Complementary, wearable biosensors capable of detecting physiological indicators of stress can enable personalized, context-aware interventions, such as relaxation prompts or breathing exercises, to improve emotional regulation in high-stress environments [28, 29]. Additionally, digital patient-facing dashboards that provide up-to-date information on wait times, treatment timelines, and care team availability may reduce uncertainty and enhance perceived transparency, thereby reducing frustration and improving the overall sense of control and satisfaction [30–32]. Collectively, these technologies offer scalable solutions to alleviate the psychological burden of hospitalization. The translation of these insights into clinical practice requires the development of assistive tools that align with patient-reported needs and preferences.

The translation of these findings into clinical practice will entail the co-design of intelligent assistive technologies that reflect patient-reported needs and emotional experiences. The integration of our survey findings into the design of intelligent assistive devices offers a promising pathway to enhance patient experiences during hospitalization. The results of this survey indicate a moderate negative correlation between prolonged waiting times and reduced mental well-being. This suggests the need for real-time updates on wait periods and interactive content to reduce anxiety [14, 26]. Additionally, communication gaps, particularly during wait times, highlight the potential of AI-driven chatbots that provide information and multi-language support to improve patient-staff interactions.One potential application of these proposed assistive technologies is an avatar system capable of providing patients with general information and entertainment during waiting periods. The implementation of such a system is feasible through the integration of specialized devices within the general waiting areas of the hospital. A more sophisticated system could be distributed at the commencement of hospitalization, subsequently providing the patient with personalized support throughout their stay.

The survey substantiates established stressors in the hospital context while embedding them in the specific context of radiological procedures. Future research should focus on co-designing these solutions with patients and clinicians to ensure usability, security, and seamless integration into hospital systems [18]. To ensure successful implementation, future research should prioritize co-design with patients and clinical staff, emphasizing emotional needs, usability, and seamless integration into hospital workflows.

### Strengths and Limitations

A key strength of this survey is its successful execution of data collection in a restricted-access clinical setting, conducted without disrupting established workflows. By preserving the integrity of routine workflows, the research provides a strong foundation for future investigations into patient well-being. A key strength of this survey is its ability to capture patients’ subjective experiences and preferences, offering insights into the psychological and emotional challenges they face in clinical environments.

However, the survey has several limitations. Its single-center design restricts the generalizability of the findings. Additionally, not all patients in the waiting area were physically or cognitively able to participate, introducing a potential selection bias. While convenience sampling was the most feasible approach given the clinical constraints, it inherently limits randomization, which may affect external validity. The sample was also predominantly male (81%), further limiting the applicability of findings across diverse patient populations. Furthermore, the limited overall sample size (N=51) restricts statistical power and constrains the ability to perform robust subgroup analyses, particularly regarding gender-based differences in emotional well-being. The pronounced gender imbalance, with only 18% female participants, reinforces the necessity of more representative sampling strategies in future studies. Expanding the sample to include more diverse patient demographics would enable a more nuanced understanding of emotional needs and technological preferences, thus enhancing the generalizability and clinical relevance of future findings. Furthermore, the survey is not equipped to determine the most effective interaction method between assistive technology and patients. This is due to the fact that the questions in the questionnaire exclusively focused on the emotional impact of hospitalization. It is evident that further research is required to ascertain the most effective methods of providing support to elderly individuals and patients who possess limited communication skills or language proficiency. Furthermore, as all responses were based on subjective perceptions, individual variability limits the generalizability of conclusions.

## Conclusion

The survey results show that prolonged delays, communication challenges, and the psychological distress of an unfamiliar hospital environment significantly affect patients’ mental well-being, resulting in a variety of emotional responses. These observations highlight clear opportunities for intelligent assistive devices to enhance mental support in clinical settings. Intelligent digital platforms have the potential to deliver customized coping strategies to alleviate the stress associated with hospital unfamiliarity, while AI-driven systems such as chatbots may improve communication by offering immediate, multilingual access to relevant information. Sensor-integrated devices provide the potential to further extend these capabilities by enabling real-time detection of emotional distress, supporting timely interventions and proactive adjustments to care processes, including those addressing stress related to wait times. The survey results indicate the potential of intelligent assistive technologies to cultivate a more responsive hospital environment, thereby establishing the foundation for future research, particularly in the field of technology usage among patient groups with communication limitations and elderly people.

## Author Contributions

AM: Conceptualization, Methodology, Investigation, Data Curation, Formal Analysis, Visualization, Writing – Original Draft; NR: Conceptualization, Supervision, Writing – Review & Editing; MV: Conceptualization, Supervision, Writing – Review & Editing

## Acknowledgments

The researchers declare no financial support influenced the design, execution, or interpretation of this research. AM gratefully acknowledges the support of the Albert und Anneliese Konanz-Stiftung through a personal *Promotionsstipendium* (doctoral scholarship).

We acknowledge the use of Generative AI (OpenAI’s GPT-4o via bwGPT [33]) for textual assistance in refining the language and presentation of this manuscript [34]. No AI-generated content contributed to the conceptualization, design, analysis, or interpretation of the research. All scientific content, including research design, methodology, analysis, and conclusions, was developed independently by the authors.

The authors have declared that no competing interests exist.

## Data Availability

The datasets generated and analyzed during the current study are available in the Zenodo repository, DOI: 10.5281/zenodo.15772899.

